# CohortDiagnostics: phenotype evaluation across a network of observational data sources using population-level characterization

**DOI:** 10.1101/2023.06.28.23291982

**Authors:** Gowtham A. Rao, Azza Shoaibi, Rupa Makadia, Jill Hardin, Joel Swerdel, James Weaver, Erica A Voss, Mitchell M. Conover, Stephen Fortin, Anthony G. Sena, Chris Knoll, Nigel Hughes, James P. Gilbert, Clair Blacketer, Alan Andryc, Frank DeFalco, Anthony Molinaro, Jenna Reps, Martijn J Schuemie, Patrick B Ryan

## Abstract

**Objective:** This paper introduces a novel framework for evaluating phenotype algorithms (PAs) using the open-source tool, Cohort Diagnostics.

**Materials and Methods:** The method is based on several diagnostic criteria to evaluate a patient cohort returned by a PA. Diagnostics include estimates of incidence rate, index date entry code breakdown, and prevalence of all observed clinical events prior to, on, and after index date. We test our framework by evaluating one PA for systemic lupus erythematosus (SLE) and two PAs for Alzheimer’s disease (AD) across 10 different observational data sources.

**Results:** By utilizing CohortDiagnostics, we found that the population-level characteristics of individuals in the cohort of SLE closely matched the disease’s anticipated clinical profile. Specifically, the incidence rate of SLE was consistently higher in occurrence among females. Moreover, expected clinical events like laboratory tests, treatments, and repeated diagnoses were also observed. For AD, although one PA identified considerably fewer patients, absence of notable differences in clinical characteristics between the two cohorts suggested similar specificity.

**Discussion:** We provide a practical and data-driven approach to evaluate PAs, using two clinical diseases as examples, across a network of OMOP data sources. Cohort Diagnostics can ensure the subjects identified by a specific PA align with those intended for inclusion in a research study.

**Conclusion:** Diagnostics based on large-scale population-level characterization can offer insights into the misclassification errors of PAs.

## BACKGROUND AND SIGNIFICANCE

Phenotype algorithms (PA) are computerized queries used to identify specific clinical events on health data sources such as electronic health records or administrative claims.[1-4] However, the reliability of evidence generated from observational studies may be threatened by misclassification errors.[5] A reproducible framework is needed to systematically evaluate PA’s for the detection, quantification, and reduction of such misclassification errors.

Misclassification errors can be assessed using metrics such as sensitivity, specificity, positive predictive value (PPV), and negative predictive value (NPV). These metrics typical depend on comparison to a gold standard reference classifier, such as a comprehensive disease registry or medical record reviews. Unfortunately, disease registries are not always available, and even when they are, they often cover only a limited range of conditions and might be incomplete.[6] Medical record reviews, while valuable, are resource-intensive, time-consuming, prone to interobserver bias, and unfeasible in large deidentified data sources.[7, 8] Furthermore, most medical record reviews provide only PPV information.

Recent advances have led to the introduction of scalable alternatives like CALIBER and PheValuator.[9, 10] Although these novel methods report on the existence and magnitude of measurement errors, they do not identify the sources of these errors or suggest modifications to the PA to enhance its performance.

This manuscript proposes a framework to address these gaps and supplements existing methods for PA evaluation.

### OBJECTIVE

In this work, we introduce a new framework to assess potential misclassification errors in PAs using population-level characterization. This framework has been integrated into CohortDiagnostics, an open-source software that is able to run on person level health data in Observational Medical Outcomes Partnership (OMOP) common data model format.[11] To illustrate its effectiveness, we apply this methodology to two distinct health conditions represented as computable phenotypes: Systemic Lupus Erythematosus (SLE) and Alzheimer’s Disease (AD).

## MATERIALS AND METHODS

### Overview

We use a data-driven approach to evaluate PAs. This method is based on a set of summary statistics (characterization of the cohort) that serve as diagnostic indicators. Each of these ‘diagnostics’ provides insights on potential misclassification errors.

To clarify, when a PA is run against a data source, the result is a ‘cohort’. A cohort is a set of individuals who satisfy all the criteria specified in the PA for a duration of time represented by cohort_start_date and cohort_end_date. ‘CohortDiagnostics’ is an open-source software tool that generates and visualizes summary statistics called diagnostics. These diagnostics include estimates of incidence rate; the breakdown of entry event codes on the index date (ie, cohort entry); the distribution of type of visits prior to, on, and after the index date; and the prevalence of all observed clinical events prior to, on, and after the index date.

Table 1 lists the entire set of diagnostics that are available in CohortDiagnostics and provides a guide on how to use it.

**Table 1:**
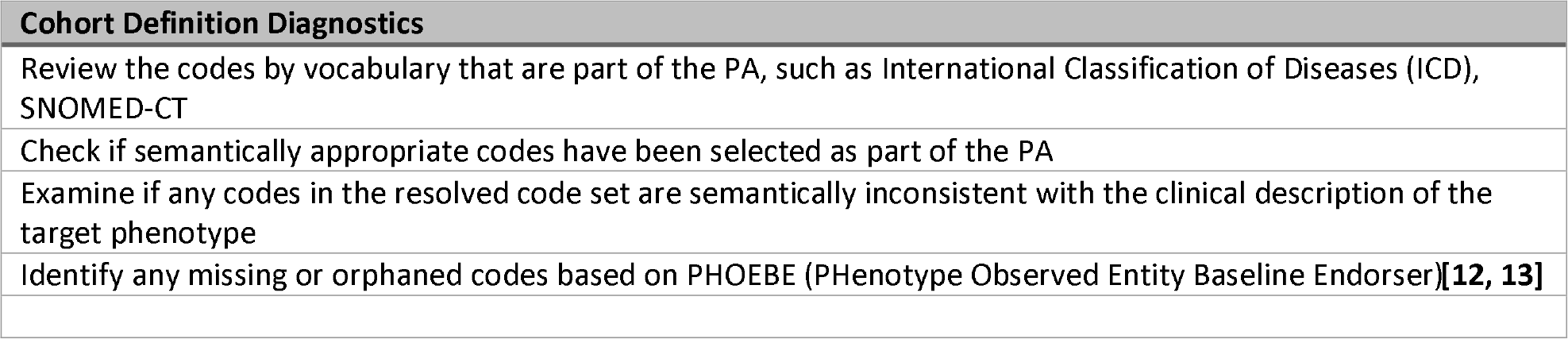
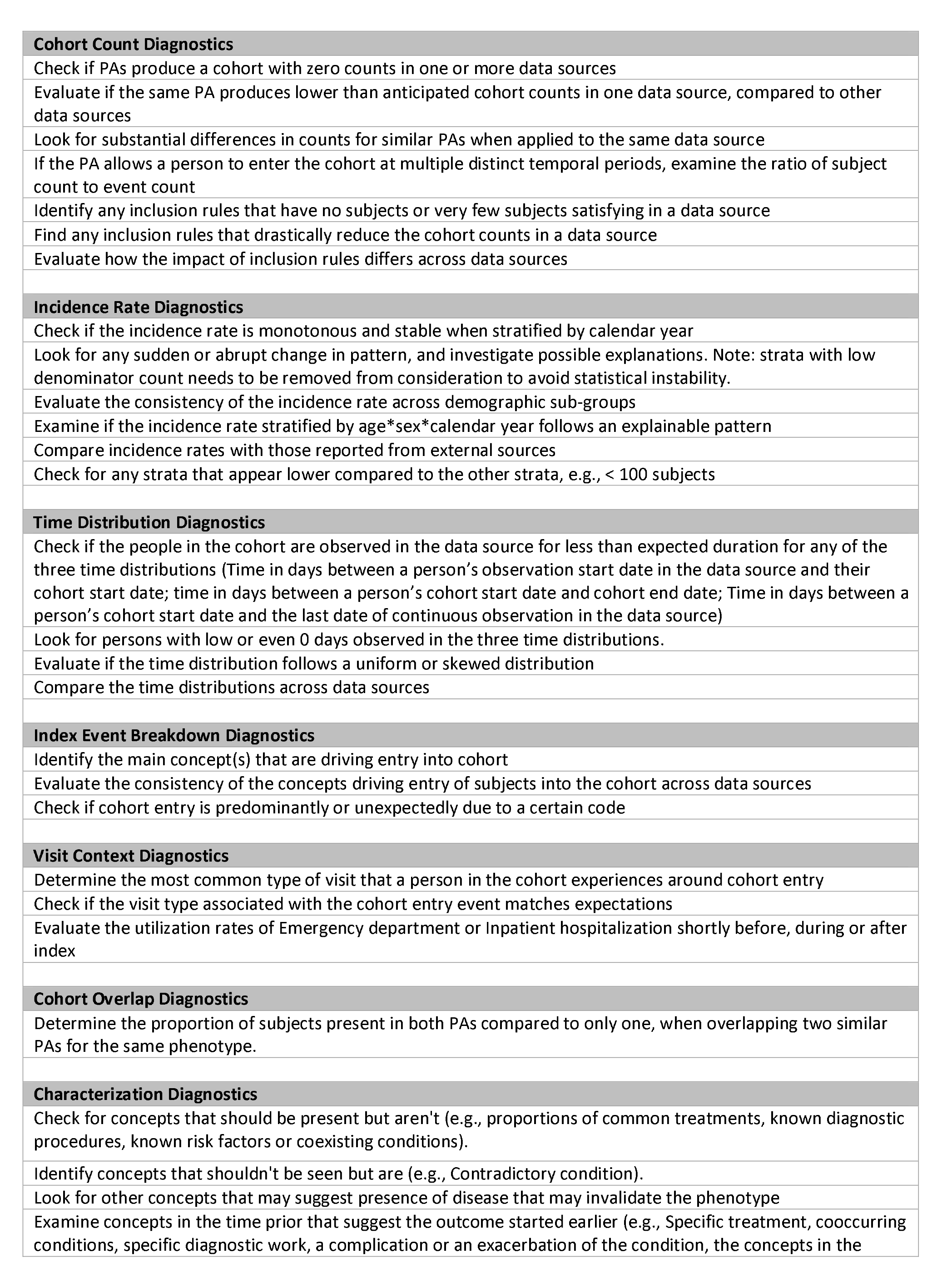
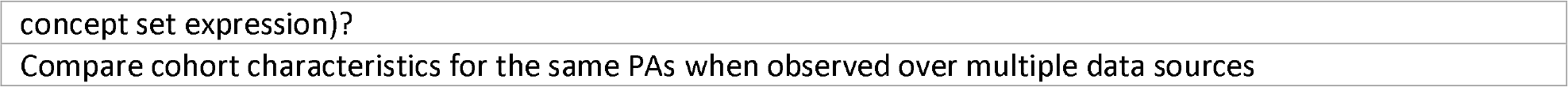
*Diagnostics and Guide on using diagnostics to infer misclassification error.*

### Incidence rates

Incidence rates are calculated for all permutations of 10-year age groups, sex, and calendar year strata. The rates are determined by dividing the number of individuals entering a stratum for the first time, termed ‘incident cohort entry’, by the sum of person-years contributed by all eligible individuals in data source who could potentially enter the strata for the first time, referred to as ‘eligible for incident cohort entry’.

Typically, incidence rates stratified by calendar year are anticipated to be present in a continuous, unvarying monotonous temporal pattern with no abrupt shifts. Should any interruptions in this pattern be observed, it could suggest alterations in either clinical practices or data capture processes, leading to potential inconsistencies in PA performance. Moreover, these incidence rate findings can be cross-referenced with expected epidemiological trends documented in the existing literature for additional validation.

### Index event breakdown

Index event breakdown shows the count of cohort entries where a specific code in the PA’s entry event criteria, coincided with the index date of cohort entry. In other words, these are codes that likely triggered the cohort entry. The frequency of these codes allows us to assess their individual contributions to the cohort.

If most individuals are entering the cohort based on a limited number of the total specified codes in the PA, this could potentially point towards specificity errors. A higher occurrence of codes that might be semantically narrower compared to the clinical definition of the phenotype may suggest sensitivity errors. Additionally, any variations in the rank order of codes among different data sources might indicate measurement heterogeneity.

### Visit Context

This diagnostic presents the count of individuals who experienced different types of healthcare visits (outpatient, inpatient, emergency department) in relation to the index date, as follows: 1) ‘Before’ represents visits that concluded within 30 days prior to the index date. 2) ’During’ accounts for visits that began before and extended up to or beyond the index date. 3) ’Simultaneous’ covers visits that initiated on the index date. 4) ’After’ includes visits that commenced within 30 days post the index date.

We anticipate that certain types of visits will be more common for specific patient phenotypes. For instance, severe acute conditions requiring intensive care will probably result in inpatient visits. A preponderance of unanticipated visit types might suggest a specificity error.

### Cohort Overlap

The cohort overlap diagnostics conducts pairwise comparison of cohorts from two PAs, reporting the individuals identified by either one or both PAs, as well as only one PA. This diagnostics in CohortDiagnostics is visualized using a Venn diagram or a table. Examining the overlap between two different PAs representing the same disease can provide insights into the potential sensitivity loss associated with one algorithm compared to the other.

### Cohort Characterization

Cohort characterization diagnostics provides a overview of the cohort using descriptive statistics on demographic factors, condition, drug exposures, measurements, and occurrences of procedure codes. For each selected data source, CohortDiagnostics displays the prevalence of all observed clinical events (denoted by codes) at different time periods relative to the index date. Default time windows include a) 365 to 31 days prior to the index date, b) 30 to 1 day before the index date, c) on the index date, d) 1 to 30 days post the index date, and e) 31 to 365 days post the index date.

Clinical events are represented through one or more codes. The prevalence is given for each code individually, and some are grouped using a vocabulary hierarchy. This diagnostic feature allows us to simulate, at the cohort level, the process by which clinicians establish and confirm clinical diagnoses. We expect individuals diagnosed with a certain disease to exhibit its signs and symptoms on or before the index date. Similarly, we anticipate diagnostic tests related to the disease to occur on or before its onset, followed by relevant treatment occurrences on or after onset. A lack of such expected characteristics might point to misclassification errors. To enable comparative analyses across multiple PAs, the tool carries out pairwise comparisons of all observed characteristics for each assessed PA. The results, including proportions or means and the standardized (mean) difference for each covariate, are presented in tables and scatter plots.

### Application

Our evaluation of PAs focuses on two distinct scenarios: First, a researcher may examine a single PA on its own merit, by looking for possible misclassification errors across one or more data sources. This evaluation could guide the researcher in adjusting the PA in successive iterations to reduce possible misclassification errors. Second, a researcher may compare the diagnostic performance of two or more PAs that represent the same clinical concept in the same data source. This would help the researcher infer which PA might has lower misclassification errors, allowing them to choose from the two the PA that offers the best performance.

To illustrate these two scenarios, we implemented our proposed framework on two clinical concepts of interest - SLE and AD. Before evaluating the PA, we ensure we understand the known clinical profile of persons we are attempting to capture in the data source. This is done by writing a clinical description for medical condition/disease, with elements like overview, presentation, diagnostic evaluation, therapy plan, risk factors, and prognosis. The authored clinical description serves as a tool that enables documentation of the shared understanding among researchers of the target clinical idea. It also provides justification for the phenotype development design choices and expected clinical attributes to look for during phenotype evaluation.

### Phenotypes

#### System Lupus Erythematous

SLE is an autoimmune disease with a wide range of severity characterized by periods of exacerbation and relative quiescence and occurs predominantly among women of child-bearing age (15 to 44 years). Based on SLE clinical description, we developed a PA that allows patients to enter the cohort on the earliest of either a diagnosis code, treatment for (ie, hydroxychloroquine, steroids, biologics, or immunosuppressants) or signs and symptoms related to SLE (ie, Inflammatory dermatosis, rash, joint or back pain, endocarditis), as long as there was at least one diagnosis code for SLE within 0 to 90 days from the entry date. All patients were required to have at least 365 days of continuous observation prior to the index date. The full PA for SLE including condition and drug codes and temporal logic is in Appendix 1.

#### Alzheimer’s Disease

AD is an age associated progressive neurodegenerative disorder and the most common cause of dementia.[14] For AD, we constructed 2 PAs. The first AD PA (referred to as the simple PA) allows patients to enter the cohort on first occurrence of an AD diagnosis. The second AD cohort is a more restrictive and is derived from the work by Imfeld et. al.,[15] requiring one of 3 inclusion criteria: 1) the first occurrence of AD diagnosis as the entry event criterion, and any of the following inclusion criteria in relation to entry date: a) a prescription on or after for AD drug, b) a second AD diagnosis any time after, c) a prior dementia test, d) a prior, simultaneous, or subsequent dementia symptom, or e) if the first occurrence was diagnosed in an inpatient setting; or having the 2) first occurrence of dementia followed by at least 2 prescriptions for AD drugs; or 3) prescription for AD drugs followed by a diagnosis of AD. Individuals were excluded if they were under 18 years of age at cohort start date, were subsequently diagnosed with diseases that, when present, make the diagnosis of AD less likely (eg, Vascular dementia, Lewy Body disease, Pick’s disease), or had an occurrence of a stroke diagnosis within 2 years before index date.

### Data

The data sources used in the evaluation are described in Appendix 2. We included 6 claims based data; JMDC, Merative^TM^ MarketScan^®^ Commercial Claims and Encounters Database (CCAE), Merative^TM^ MarketScan^®^ Medicare Supplemental and Coordination of Benefits Database (MDCR), Merative^TM^ MarketScan^®^ Multi-State Medicaid Database (MDCD), IQVIA^®^ Adjudicated Health Plan Claims Data (Pharmetrics Plus), Optum’s Clinformatics^®^ Data Mart - Socio-Economic Status (Optum SES) and 4 electronic medical record (EHR) data; IQVIA^®^ LPD in Australia (LPDAU), IQVIA^®^ Disease Analyzer France (France DA), IQVIA^®^ Disease Analyzer Germany (German DA), Optum^®^ de-identified Electronic Health Record dataset (Optum EHR). These data sources have been standardized to the Observational Medical Outcomes Partnership (OMOP) Common Data Model (CDM).[16, 17] Extract, transform, and load (ETL) specifications for all data sources except LPDAU, France DA, German DA, and Pharmetrics Plus are available at ETL-LambdaBuilder.[18] The standardized data are assessed using a rigorous data quality process to evaluate conformance, completeness, and plausibility of the data.[19]

### Data analysis (CohortDiagnostics software)

CohortDiagnostics is open-source software application written in the R programming language that implements the described theoretical framework.[20] Given a set of instantiated cohorts, a set of cohort definition details, and a connection to a remote database with person level data converted to the OMOP CDM [16, 17] (version 5.3+), CohortDiagnostics produces a set of aggregate summary statistics called Diagnostics. The output contains no patient-level data and has additional privacy protection using minimum cell count thresholds.[21] All output conforms to the prespecified CohortDiagnostics results data model and is formatted as unencrypted comma separated value (.csv) files (an intentional design decision to allow an investigator to audit compliance with privacy governance). The output .csv files, from one or more data sources, may then be combined and the results reviewed using an interactive R Shiny web application called DiagnosticsExplorer. The software and user documentation are available on OHDSI Github repository called CohortDiagnostics.[22]

## RESULTS

Table 2 summarizes the number of patients who met the definitions for SLE and the 2 AD PAs in each data source. Below we provide brief overviews of the key insights informed by the evaluation process. The full output of CohortDiagnostics is available in the interactive website.[23]

**Table 2:**
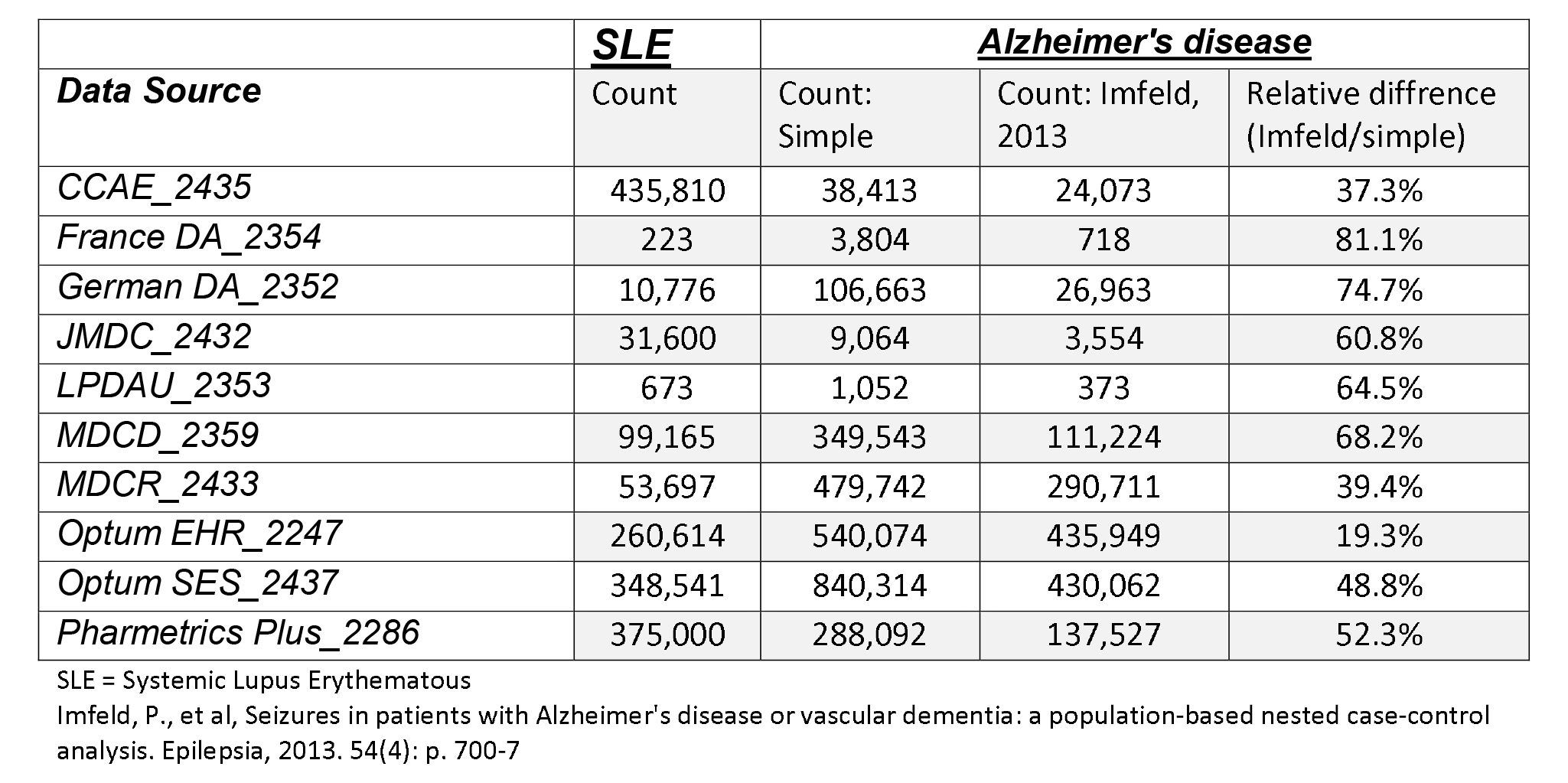
*Cohort counts for the phenotype algorithms for Systemic Lupus Erythematous and Alzheimer Disease*

### Systemic Lupus Erythematous

#### Insights from incidence rate plots

Figure 1 illustrates the pattern of incidence rate of SLE in each data source, stratified by age, gender and calendar year. Except for 2 general practitioner data sources France Disease Analyzer (France DA) and Germany Disease Analyzer (Germany DA), we observed high concordance across data sources for an incidence range from 30 to 50 per 100,000 person-years. Incidence rate estimation variation because of database heterogeneity in can be substantial, and such variation is not necessarily an evidence of measurement error.[24] However, observing concordance among data sources provides some reassurance that the PA measurement error is not causing substantial incidence rate heterogeneity.

**Figure 1:**
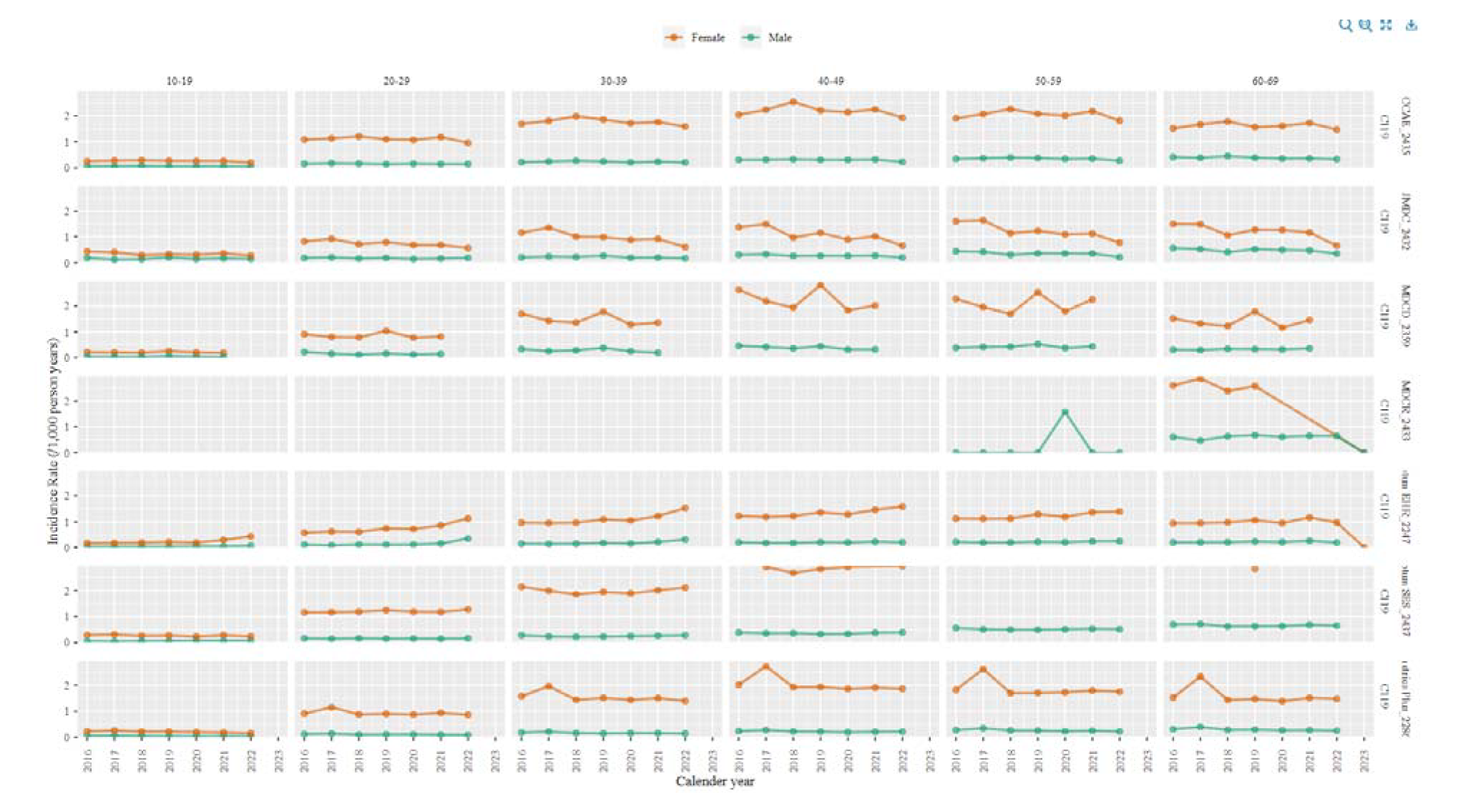
*Incidence rate of Systemic Lupus Erythematous stratified by age decile, gender, and calendar year*

As expected, females have approximately 5-fold greater incidence of SLE compared with males. However, the rates increase by age, and peak around 40 to 50 years, which is slightly older than previously reported typical age of SLE onset of 15-44 years.[25] This may imply sensitivity error among younger age patients (eg, younger women may receive treatment for SLE like symptoms without a diagnosis) or index date misclassification (eg, older patients may already have had the disease but its onset was not recorded in the data source).

#### Insights from index event breakdown

Across data sources, a substantial proportion of individuals enter the SLE cohort based on SLE symptoms or treatment. This indicates that many patients receive treatment for SLE, before their diagnosis is coded and recorded for administrative or clinical purposes. That is, a diagnosis date is observed in the data source, but this date lags the date persons could be presumed to have the disease (represented by date treatment or symptom onset). This represents index date misclassification error.

Lastly, all the events (appearing as codes) observed on the index date are related to SLE, which suggests the absence of specificity error or false positives.

#### Insights from cohort characterization

Figure 2 is a screen shot from the CohortDiagnostics tool showing the most prevalent conditions and drug exposures observed in the Optum^®^ EHR data source among the SLE cohort on the index date. SLE treatments such as prednisone, hydroxychloroquine, methotrexate and cyclophosphamide were observed on or shortly after the index date. Some individuals started these drugs in the period 365 to 30 day prior to index date, indicating potential index date misclassification. Consistent with the clinical description of SLE which stated that follow-up visits were expected, we observed SLE diagnosis codes occurring post index (30-50%). Laboratory tests such as urinalysis and antinuclear antibody were also observed (eg, in 7 to 10% in Optum^®^ EHR on index date) and these tests clustered temporally around the index date. Observing expected baseline and post index characteristics and clinical events suggests that the patients returned by the SLE PA are likely true cases and that misclassification may be limited.

**Figure 2:**
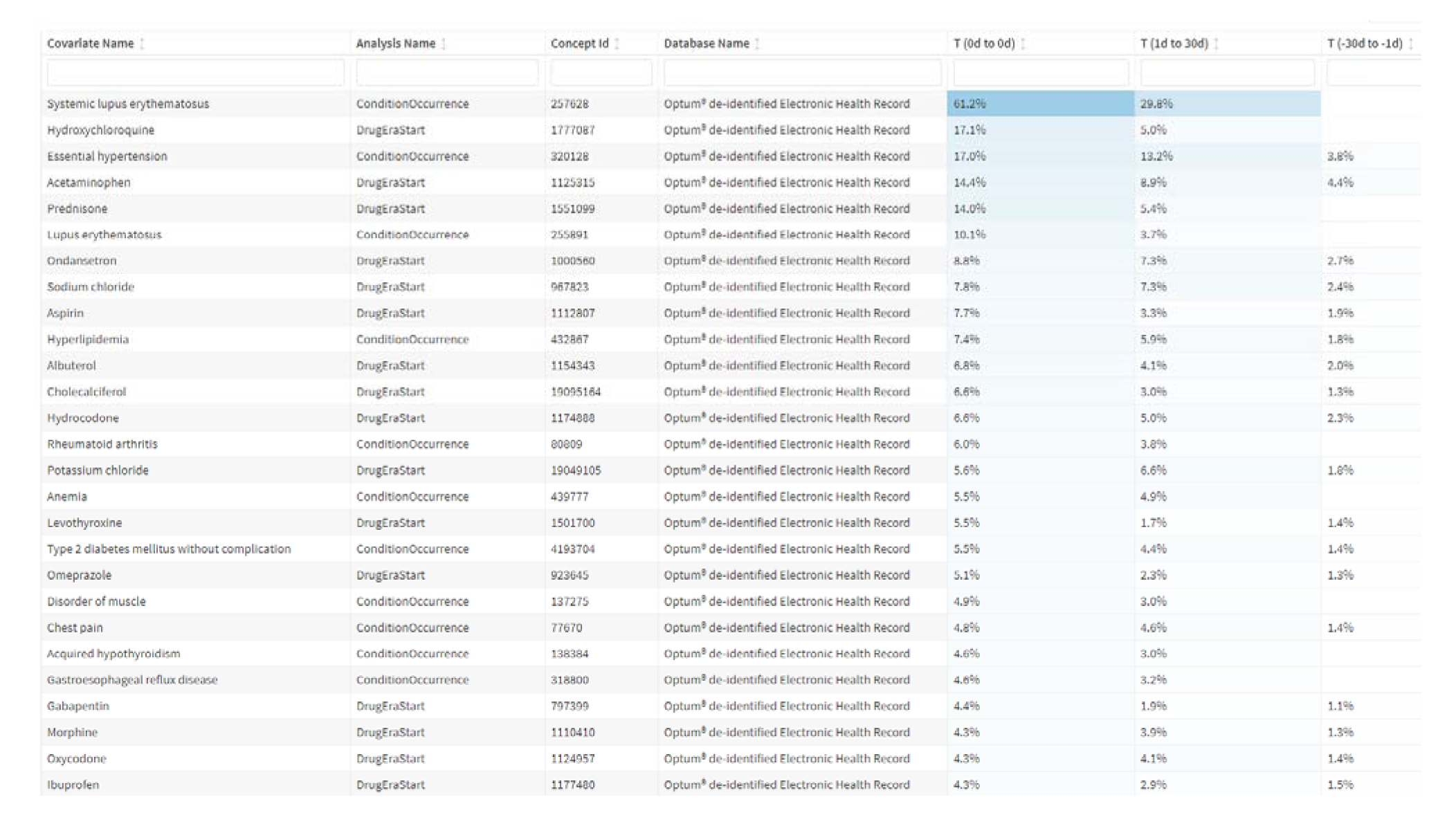
*Characterization output from CohortDiagnostics tool showing the most prevalent conditions and drug exposures on or around index date*

### Alzheimer Disease

#### Insights from cohort overlap

In all data sources, the Imfeld et.al. PA returned fewer patients (19% to 81%) compared with the simpler AD PA (See Table 1).[15] In the cohort overlap, among individuals who were present in either cohort, the proportion of individuals present in both cohorts ranged between 18% and 45%. Further, 35% to 81% of individuals were identified only by the simple PA; and 0% to 20% were identified only by the Imfeld et al PA. We can infer that the simpler PA is likely to have higher sensitivity compared with the Imfeld et al PA.

#### Insights from visit context

The distributions of the visit type around the index date among the 2 cohorts were comparable in most data sources with less than 10% of the individuals in either cohorts identified during or at the start of an inpatient visit. This suggests that neither AD PAs were likely to capture more severe cases of AD.

#### Insights from cohort characterization

Table 3 reports a selected set of characteristics from the 2 AD cohorts at baseline ie, from 365 days before the index date up to and including the index date in the Optum^®^ EHR data source (Data from all other data sources are available in the CohortDiagnostics shiny app). The covariates are defined using the Systematized Nomenclature of Medicine Clinical Terms (SNOMED-CT) vocabulary hierarchy grouping. We observe that even though the 2 cohorts were defined using different PAs and have considerably different number of patients with less than 50% overlap, the distributions of the main baseline characteristics were comparable.

**Table 3:**
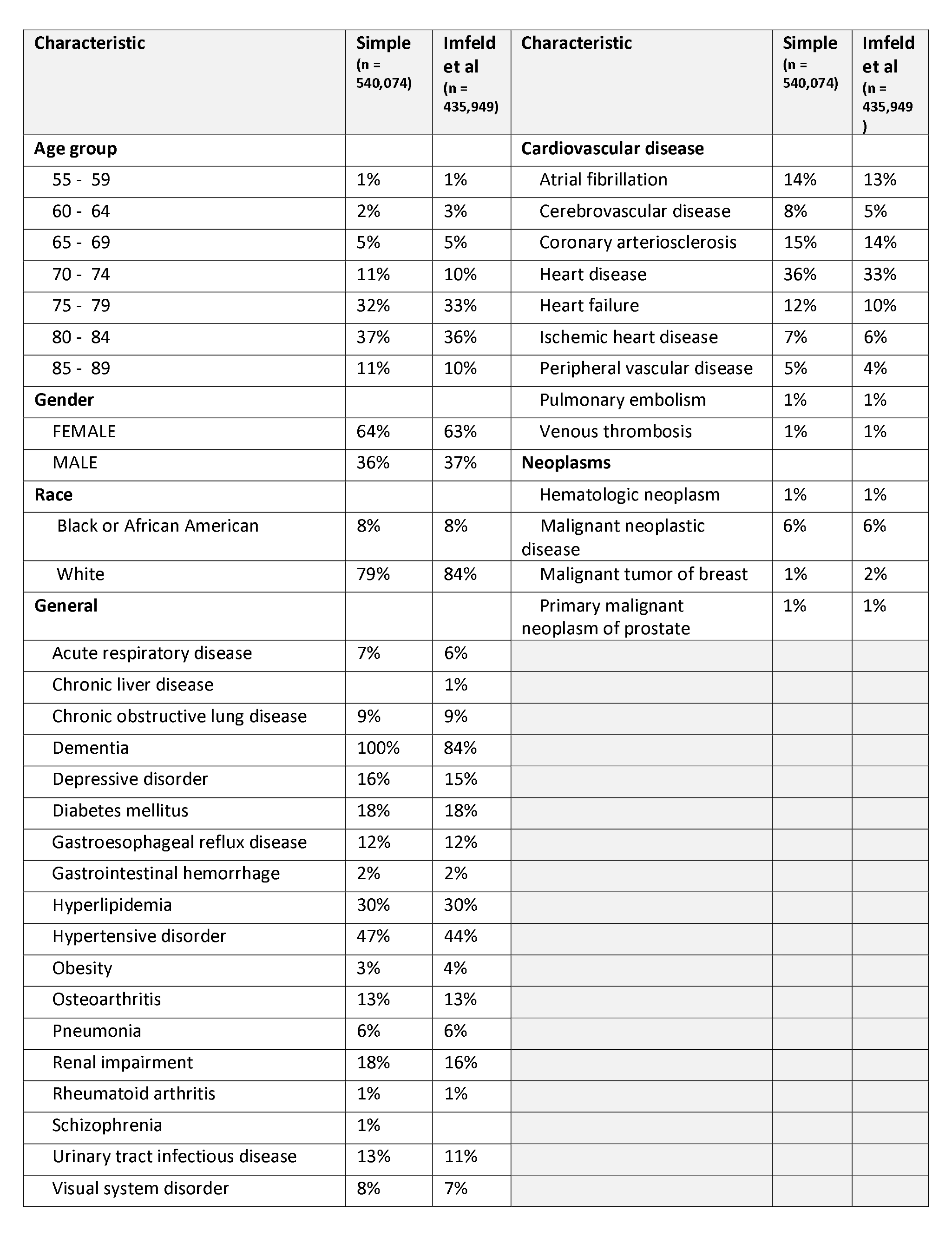
*Selected baseline characteristics among patients with Alzheimers Disease by Phenotype Algorithm.*

Figure 3 is a screenshot from CohortDiagnostics that illustrates the covariate balance between the 2 AD PAs in Optum^®^ EHR on 3 different time periods around the index date. Overall, we observed that most features near the diagonal, indicating comparable cohort characteristics distribution between the 2 cohorts. However, some covariates are off the diagonal with a larger a Standardized Mean Difference. For example, during 30 days to 1 days before index, we observed higher prevalence of vascular dementia, and other late effects of cerebrovascular accidents in the simple PA compared with the Imfeld et al PA. This suggests that the Imfeld et al PA is less likely to misclassify cerebrovascular accident events as AD. We also observed that the simpler PA had higher utilization of drugs commonly used in AD such as donepezil and memantine in the same immediate period prior to index date. Conversely, the Imfeld et al PA demonstrated higher utilization of these drugs on index. Both PAs had similar utilization after index. This suggests that the simple PA is subject to higher index date misclassification compared with the complex PA.

**Figure 3:**
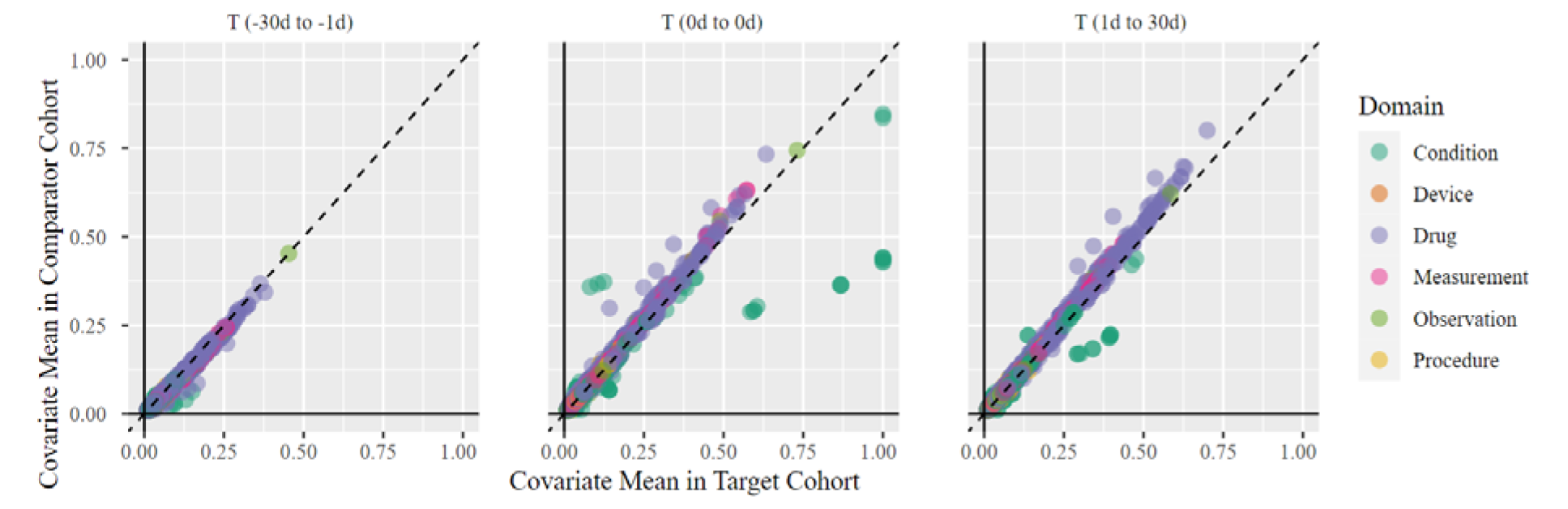
*Covariate balance between the 2 Alzheimer’s Disease Phenotype Algorithms*

When we compared covariates constructed from codes that were not part of either AD PA entry event, we observed considerable cohort similarity. This suggests that the 2 PAs identified patients with the similar clinical profiles despite incomplete cohort overlap. Overall, the descriptive data of these PAs for AD revealed that, while the Imfeld et. al. identified fewer patients (raising concerns about its sensitivity), we did not observe a higher prevalence of clinical characteristics that strongly suggest a higher specificity when compared with the other simple PA.

## DISCUSSION

We have developed and integrated an empirical methodology for the evaluation of PAs into a new tool designed for the OMOP Common Data Model called CohortDiagnostics. We have demonstrated this evaluation framework on one SLE and 2 AD PAs. Our evaluation framework categorizes errors into three types: sensitivity errors, specificity errors and index date misclassification errors, providing a consistent means of assessment. This approach allows for the identification and assessment of these errors in any PA by reviewing population-level characterization.

In our application, we conclude that the SLE PA demonstrates acceptable operating characteristics and is suitable for use across the data sources assessed, even though there is potential index event misclassification. On the other hand, we found that the Imfeld et al PA may have lower sensitivity than the simple PA. The simple PA has index event misclassification and a potential specificity error explained by the observed cerebrovascular accidents events.

We have shown that this empirical and scalable framework for PA evaluation offers insights into misclassification errors. It not only detects the existence of these errors but provides an understanding of their direction and magnitude. We demonstrate that it can provide reasons for the origin of such errors, enabling researchers to refine their PAs iteratively. This method can work together with traditional case-level retrospective medical record adjudication or innovative approaches like PheValuator, which quantify estimates of measurement error.[10, 26] When paired with validation analyses for quantifying measurement errors, our population-level characterization leads to a comprehensive understanding of a PA’s performance.

Our software tool, CohortDiagnostics, performs extensive diagnostics across multiple data sources for one or more PAs. It presents results in a privacy-compliant format. It is designed to perform phenotype evaluation across an observational database network. This feature allows a coordinator site to distribute a self-contained phenotype evaluation study package to each contributing data partner site, which can then independently execute it. After the execution, each site may share aggregate summary statistics back to the coordinator site, complying with local data governance and privacy policies. These site-level summary statistics can then be aggregated into one integrated viewer for collaborative review. This aggregated data can be used by a team of experts to discuss the merits of the PAs under evaluation and to understand associated misclassification errors. This framework has been recently implemented in numerous observational network studies and collaborations.[27-30]

The network-based phenotype evaluation process reinforces confidence in a PA. It allows for the evaluation of the consistency of diagnostics across different data sources, geographical locations, and time periods. Consistent trends in misclassification errors increase our confidence that our PAs have reliable operating characteristics, rather than representing an artifact from a specific data source. Such findings are crucial as they support the conclusion that a PA is applicable across various data sources. Moreover, evaluating a PA across a network offers valuable insights into different clinical settings, practices, and data capture processes. We are optimistic that this framework will encourage the use of more robust and externally valid PAs.

CohortDiagnostics also informs code selection during phenotype development. Selecting the right set of code to represent a clinical idea of interest is known be challenging and inconsistent.[31] While code selection should be guided by clinical judgment, the empirical impact of these judgment can be readily evaluated through our tool. This evaluation can measure the effect of alternative codes on the PA performance by assessing the impact on counts and characteristics.

Despite its strengths, our approach has some limitations. It cannot numerically quantify measurement errors and should be used in conjunction with other methods that include a gold standard, such as PheValuator, or other validation methods.[32] Furthermore, analyzing descriptive results to gain insights on misclassification errors can be subjective and time-consuming. More methodological research is required to formalize a scalable, reproducible process and establish empirically driven. Finally, this approach is based on the assumption that the evaluation data sources have been standardized to the OMOP CDM and have undergone data quality review and it is fit for research use.[19]

## CONCLUSION

In this paper, we introduce a framework for phenotype evaluation, that is intended to be done prior to observational research. It helps ensure that the individuals identified by the PA are consistent with the profiles of the patients we intend to study. Utilization of this framework enhances researchers’ confidence in the validity of their study outcomes. The framework has been integrated into the CohortDiagnostics software. We have shown how this open-source software can enable collaborative research within a broad research community and can scale to multiple PAs, over multiple data sources that can be repeated over multiple time periods enabling creation of a repository of such evaluations.[3, 4]

## DECLARATIONS

## Data Availability

The data that support the findings of this study are available to license from Merative, Optum, IQVIA and JMDC.

## Acknowledgments

None

## Ethics approval

NA

## Conflicts of interest / Competing Interests

All authors are employees of Janssen Research & Development, LLC, and shareholders of Johnson & Johnson (J&J) stock.

This study was sponsored by Janssen Research & Development, LLC.

## Author Contributions

All coauthors contributed to the conceptualization, drafting, editing, and approving the manuscript.

## Appendix 1 Systemic lupus erythematosus indexed on signs, symptoms, treatment, or diagnosis (FP)

### Human Readable Cohort Definition

#### Cohort Entry Events

People enter the cohort when observing any of the following:

1. condition occurrences of ‘SLE or signs and symptoms suggestive of SLE’; having at least 1 condition occurrence of ‘Systemic lupus erythematosus (SLE)’ for the first time in the person’s history, starting between 0 days before and 90 days after ‘SLE or signs and symptoms suggestive of SLE’ start date.
2. drug exposures of ‘SLE treatments’; having at least 1 condition occurrence of ‘Systemic lupus erythematosus (SLE)’ for the first time in the person’s history, starting between 0 days before and 90 days after ‘SLE treatments’ start date.

Limit cohort entry events to the earliest event per person.

#### Cohort Exit

The person exits the cohort at the end of continuous observation.

#### Cohort Eras

Entry events will be combined into cohort eras if they are within 0 days of each other.

### Concept Sets

#### Systemic lupus erythematosus (SLE)

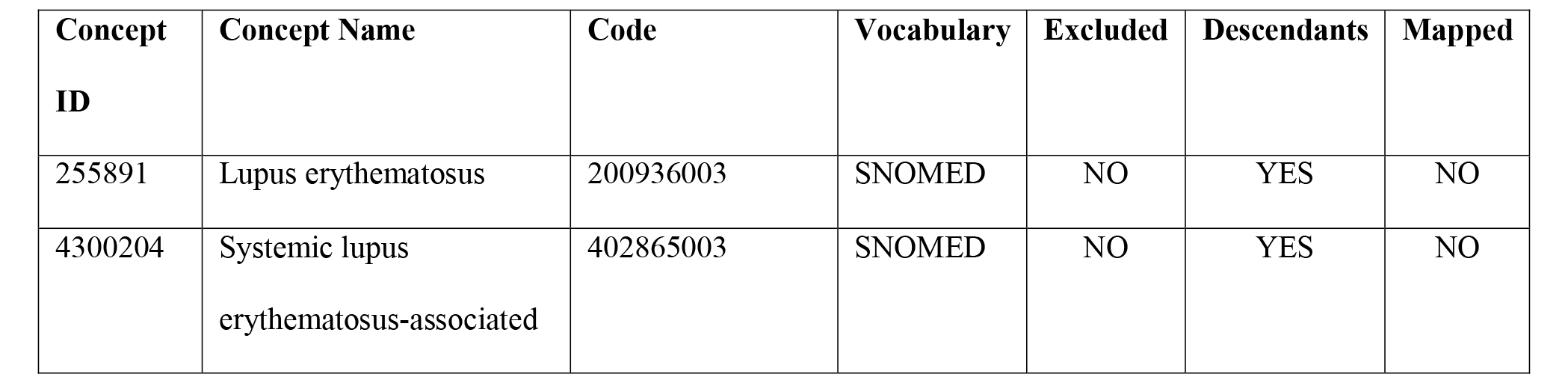
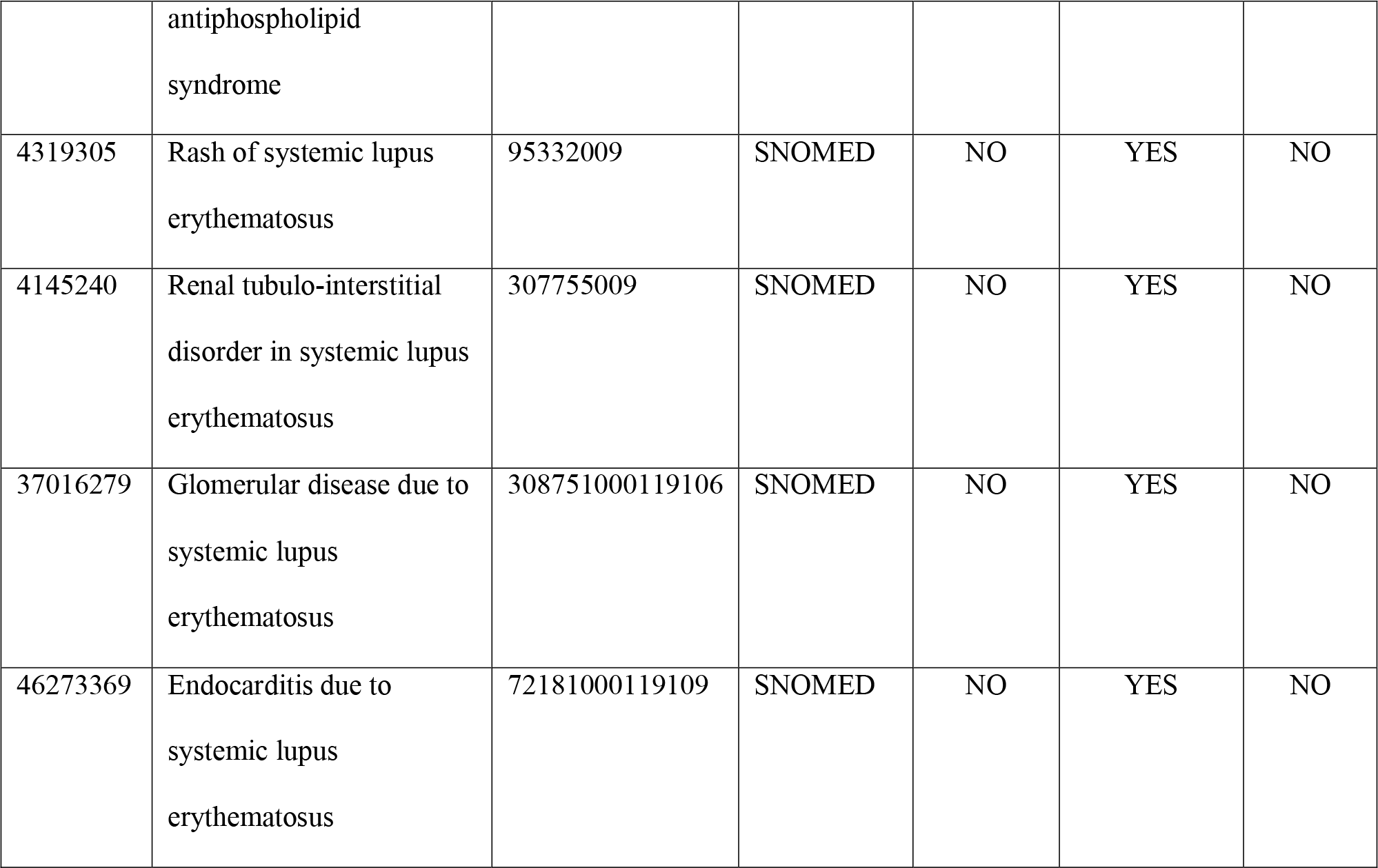

#### SLE treatments

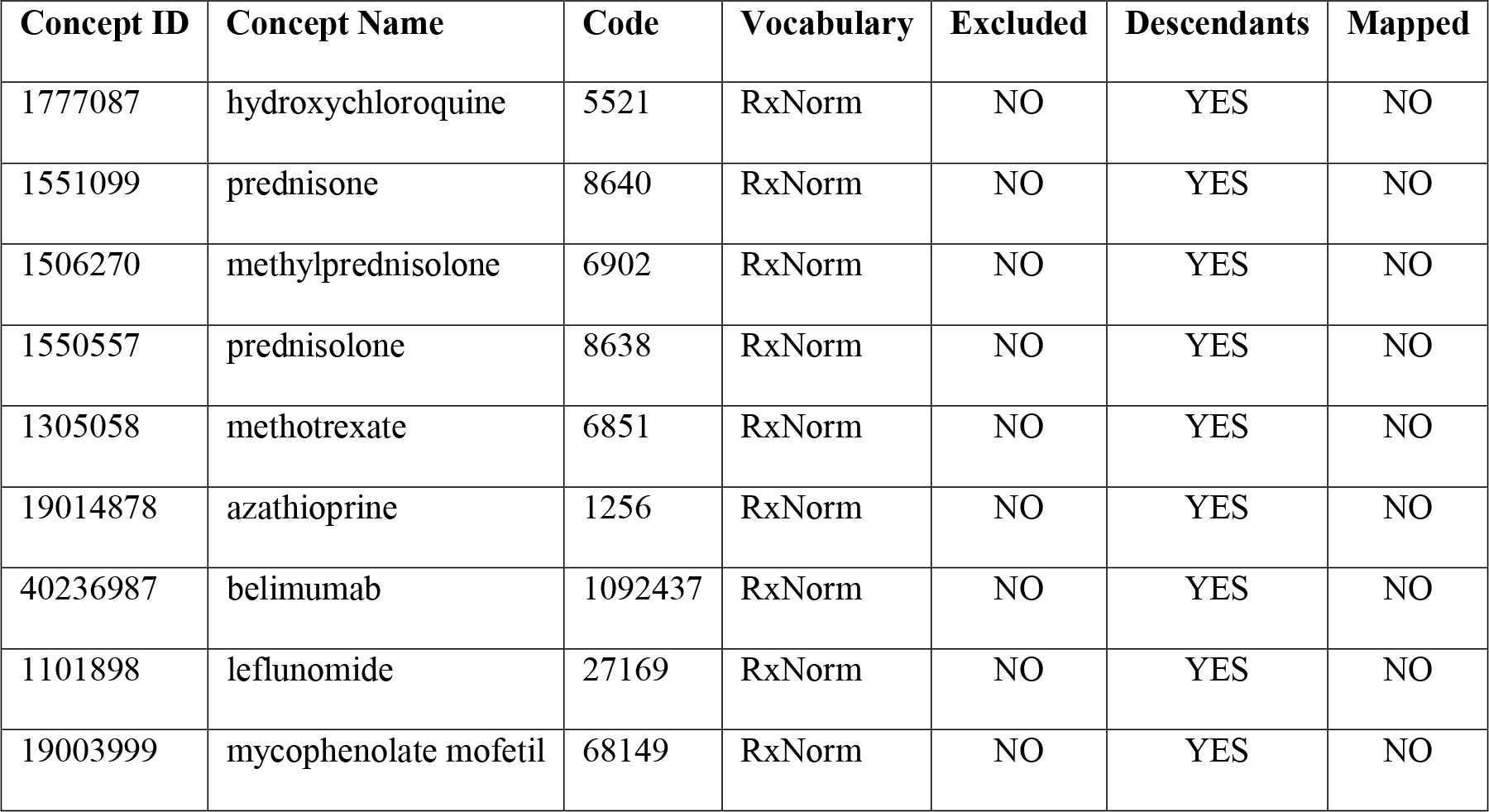

#### SLE or signs and symptoms suggestive of SLE

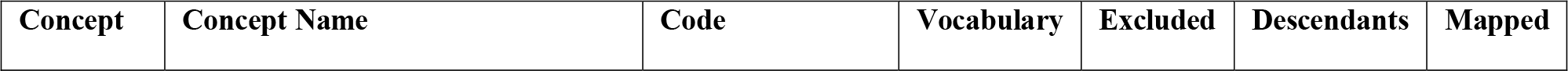
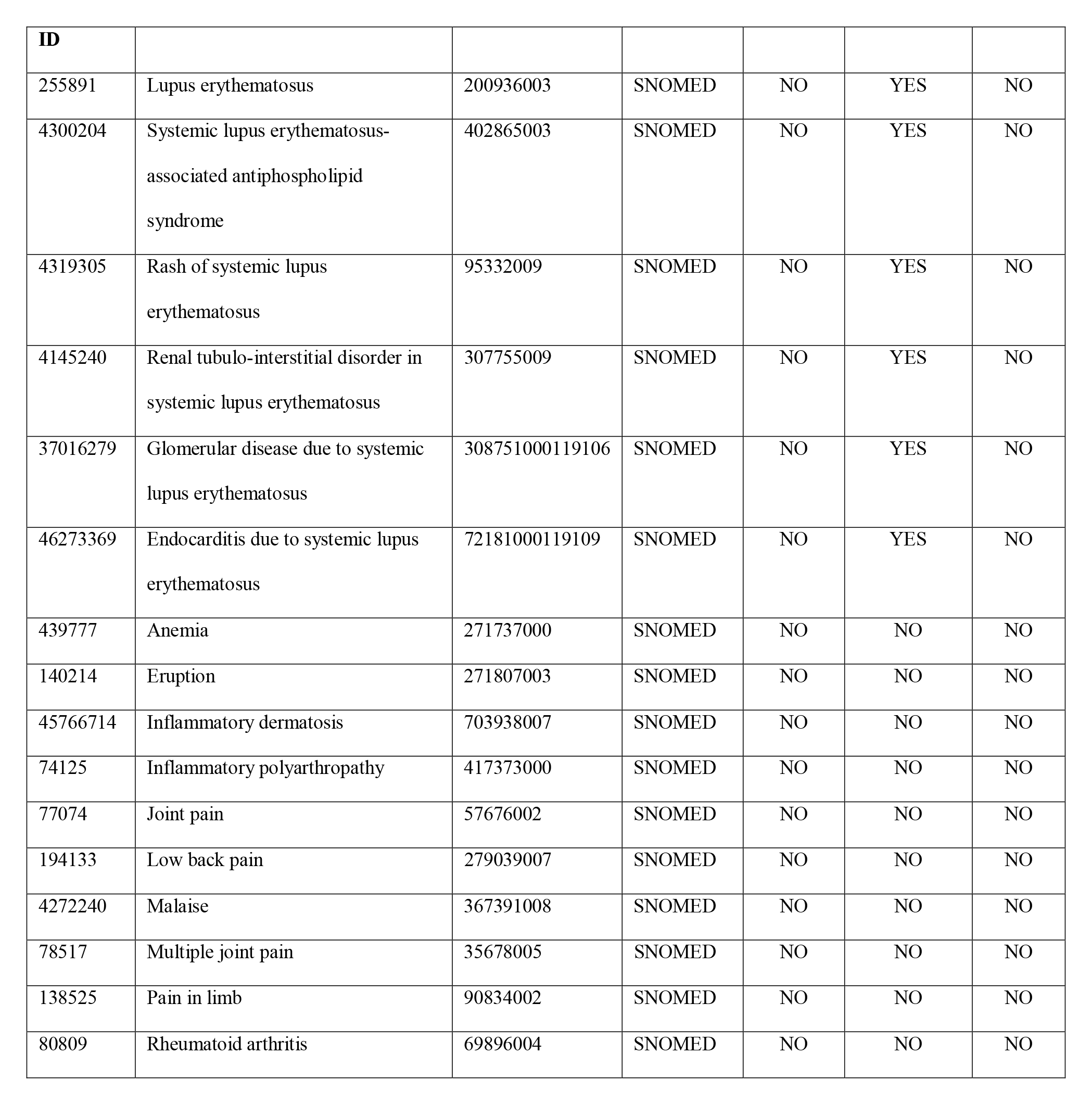

## Alzheimer’s disease (Simple)

### Human Readable Cohort Definition

#### Cohort Entry Events

People enter the cohort when observing any of the following:

1. condition occurrence of ‘Alzheimer’s disease’ for the first time in the person’s history.

Limit cohort entry events to the earliest event per person.

#### Cohort Exit

The person exits the cohort at the end of continuous observation.

#### Cohort Eras

Entry events will be combined into cohort eras if they are within 0 days of each other.

### Concept Sets

#### Alzheimer’s disease

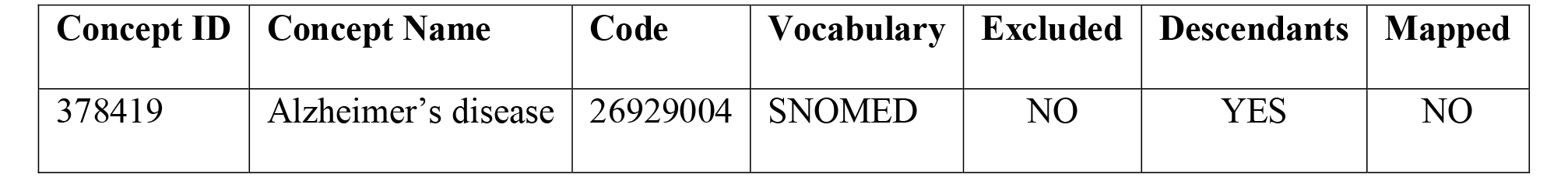

## Alzheimer’s disease (based on Imfeld, 2013)

### Human Readable Cohort Definition

#### Cohort Entry Events

People with continuous observation of 365 days before event may enter the cohort when observing any of the following:

1. condition occurrence of ’Alzheimer Disease ’ for the first time in the person’s history; with any of the following criteria:

1. having at least 1 drug exposure of ’Prescription for an Alzheimers disease drug’, starting between 0 days before and all days after ’Alzheimer Disease ’ start date.
2. having at least 1 condition occurrence of ’Alzheimer Disease ’, starting 1 days after ’Alzheimer Disease ’ start date.
3. having at least 1 procedure occurrence of ’Specific dementia test ’, starting anytime on or before ’Alzheimer Disease ’ start date.
4. having at least 1 measurement of ’Specific dementia test ’, starting anytime on or before ’Alzheimer Disease ’ start date.
5. having at least 1 observation of ’Specific dementia test ’, starting anytime on or before ’Alzheimer Disease ’ start date.
6. having at least 1 condition occurrence of ’Dementia symptoms ’.
7. having at least 1 visit occurrence of ’Inpatient or ER visit’, starting anytime on or before ’Alzheimer Disease ’ start date and ending between 0 days before and all days after ’Alzheimer Disease ’ start date.
2. condition occurrence of ’Dementia’ for the first time in the person’s history; having at least 2 drug exposures of ’Prescription for an Alzheimers disease drug’, starting between 0 days before and all days after ’Dementia’ start date.
3. drug exposure of ’Prescription for an Alzheimers disease drug’ for the first time in the person’s history; having at least 1 condition occurrence of ’Alzheimer Disease ’, starting between 0 days before and all days after ’Prescription for an Alzheimers disease drug’ start date.

Limit cohort entry events to the earliest event per person.

#### Inclusion Criteria

*1. No occurrence of any other specific dementia diagnosis (e.g., VD, Pick’s disease, or Lewy body dementia [LBD]) after the Alzheimer’s disease diagnosis date*

1. Entry events having at most 0 condition occurrences of ’Other specific dementia diagnosis (e.g., VD, Pick’s disease, or Lewy body dementia [LBD])’, starting 1 days after cohort entry start date.

*2. No occurrence of Stroke diagnosis within 2 years prior to the Alzheimer’s disease diagnosis date*

Entry events having at most 0 condition occurrences of ’Stroke (ischemic or hemorrhagic)’, starting between 730 days before and 0 days before cohort entry start date.

*3. >= 18 years old*

Entry events with the following event criteria: who are >= 18 years old.

#### Cohort Exit

The person exits the cohort at the end of continuous observation.

#### Cohort Eras

Entry events will be combined into cohort eras if they are within 0 days of each other.

### Concept Sets

#### Alzheimer Disease

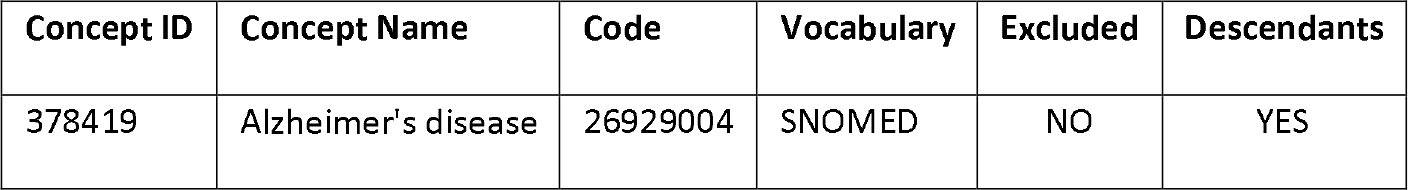

#### Dementia

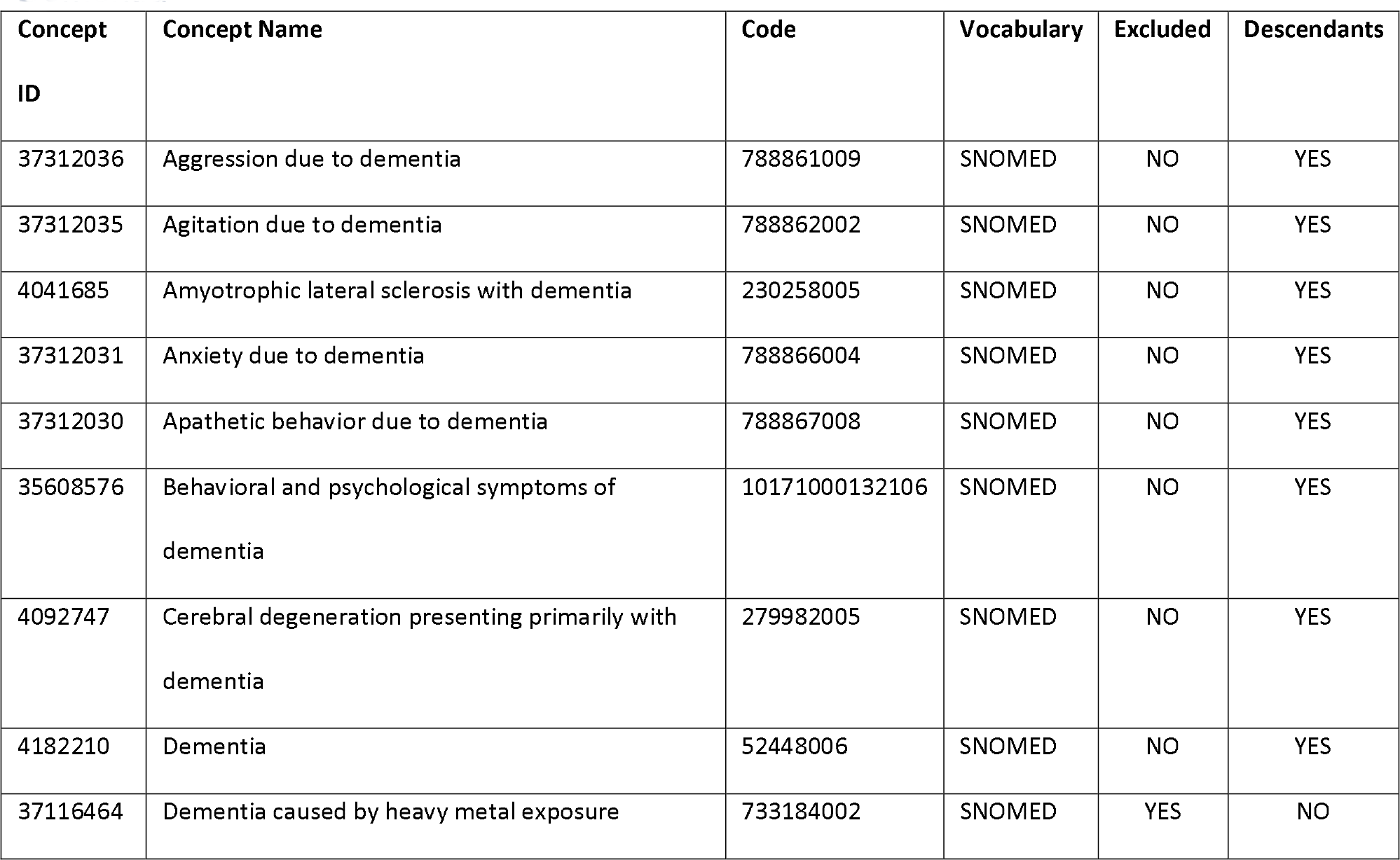
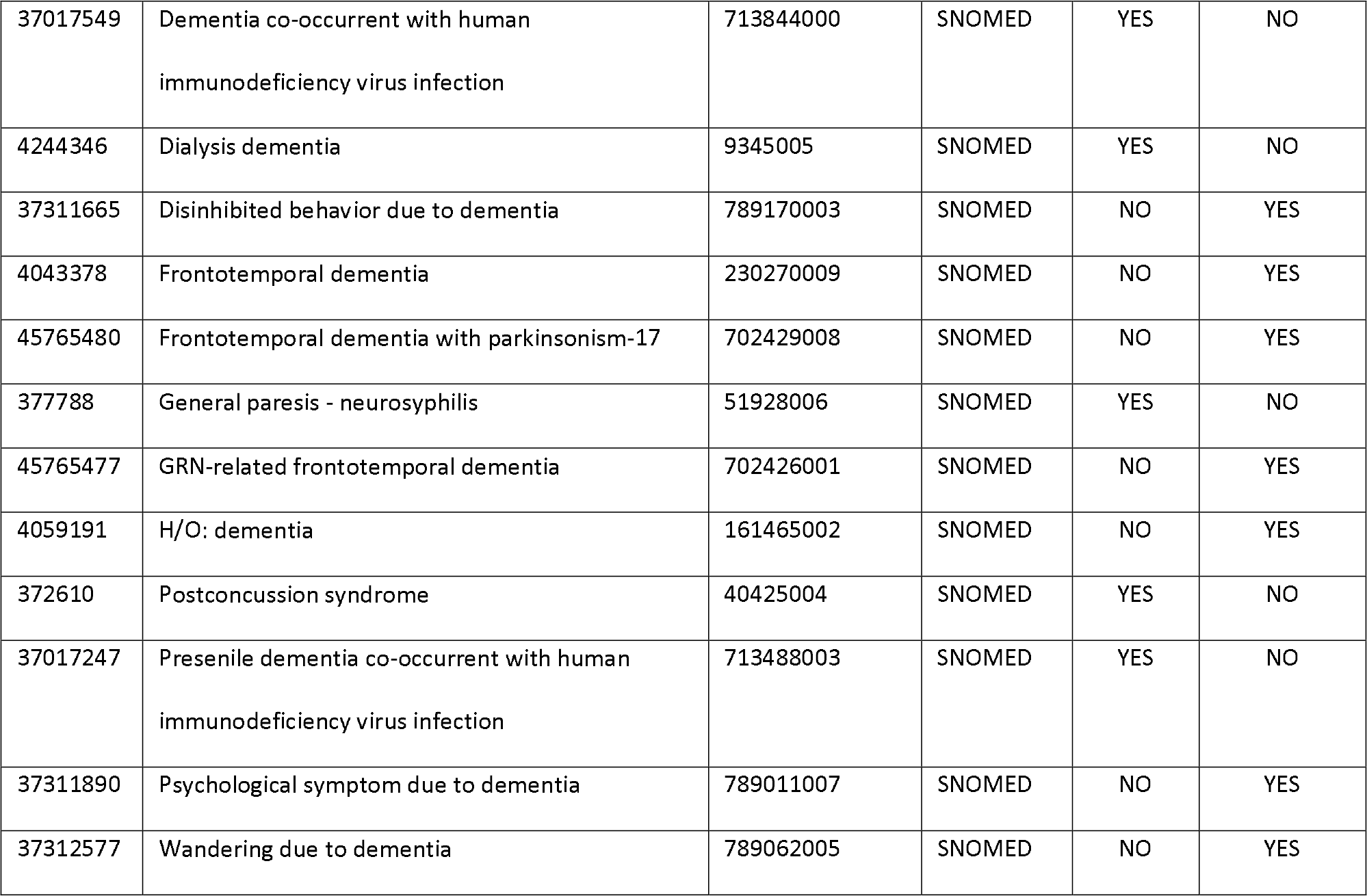

#### Prescription for an Alzheimers disease drug

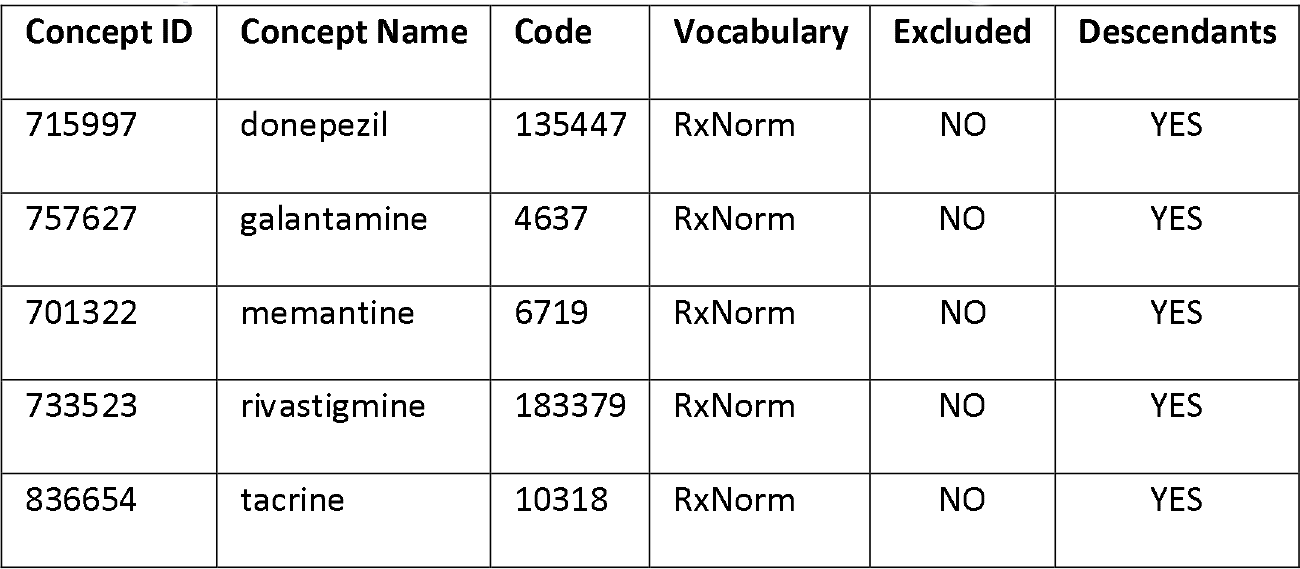

#### Specific dementia test

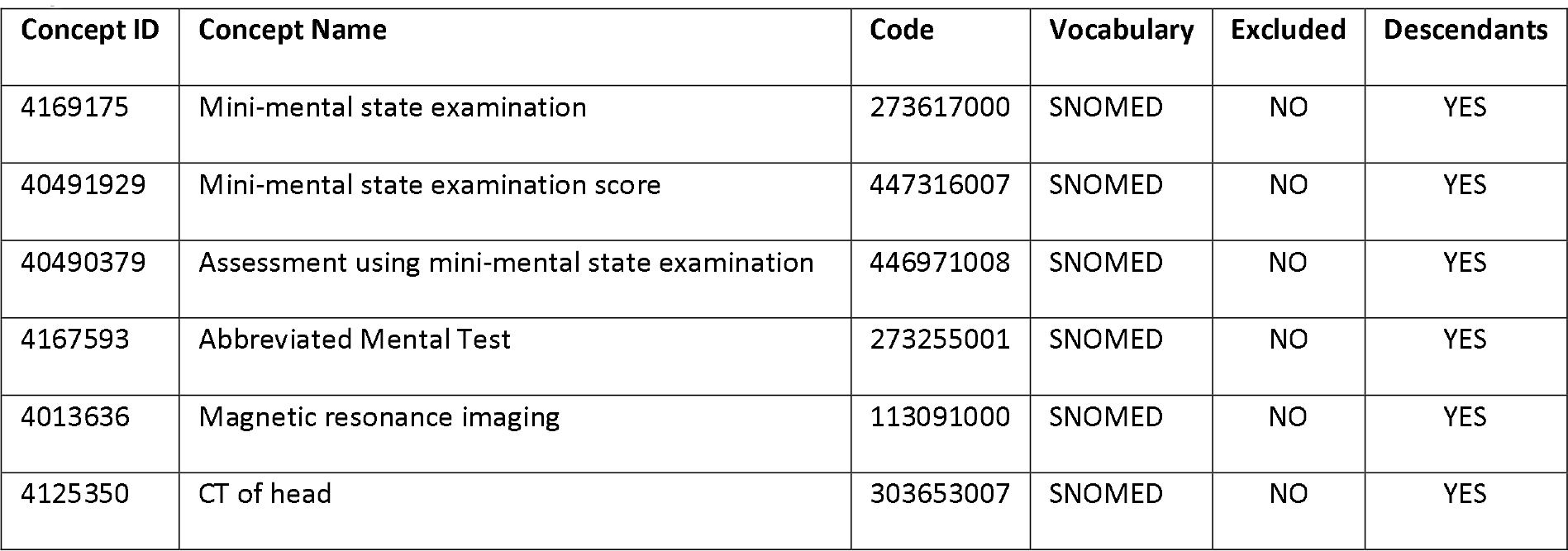
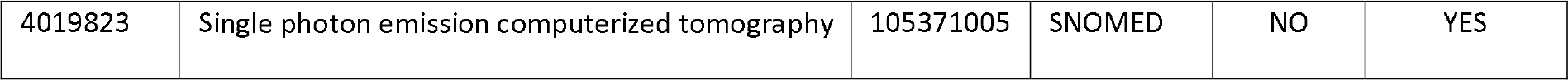

#### Dementia symptoms

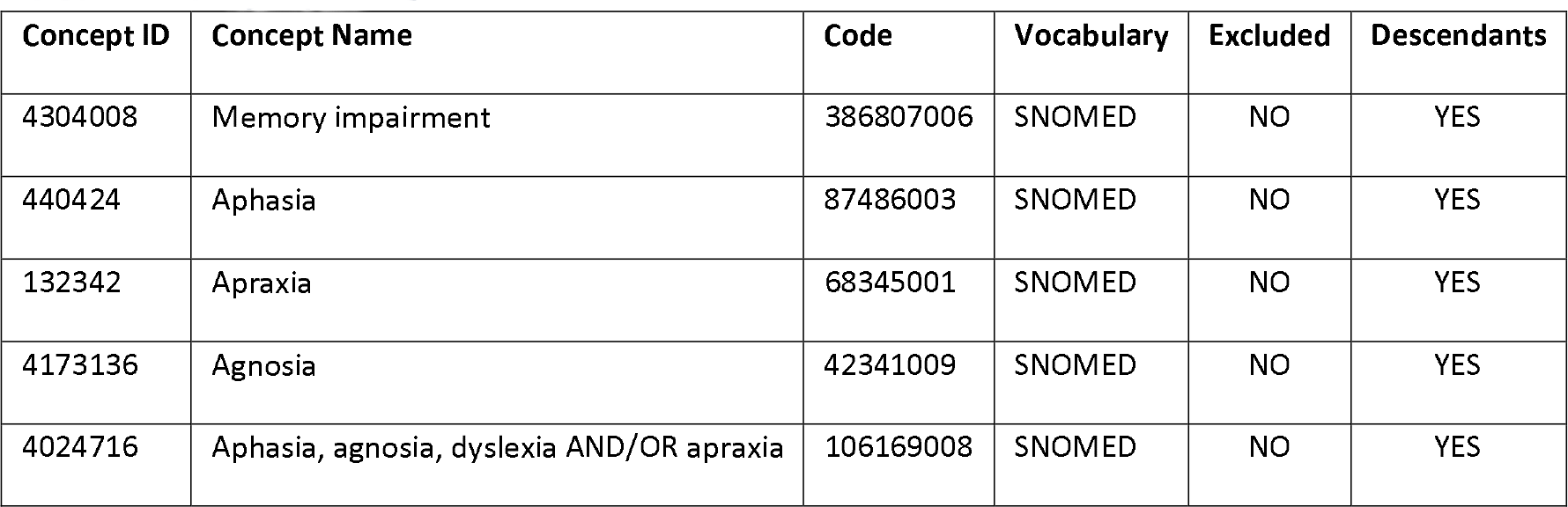

#### Other specific dementia diagnosis (e.g., VD, Pick’s disease, or Lewy body dementia [LBD])

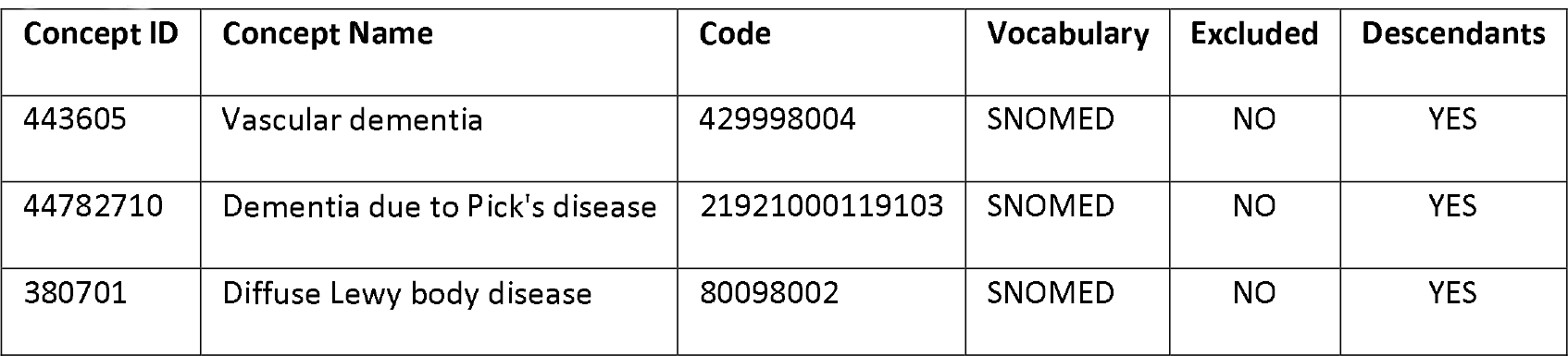

#### Stroke (ischemic or hemorrhagic)

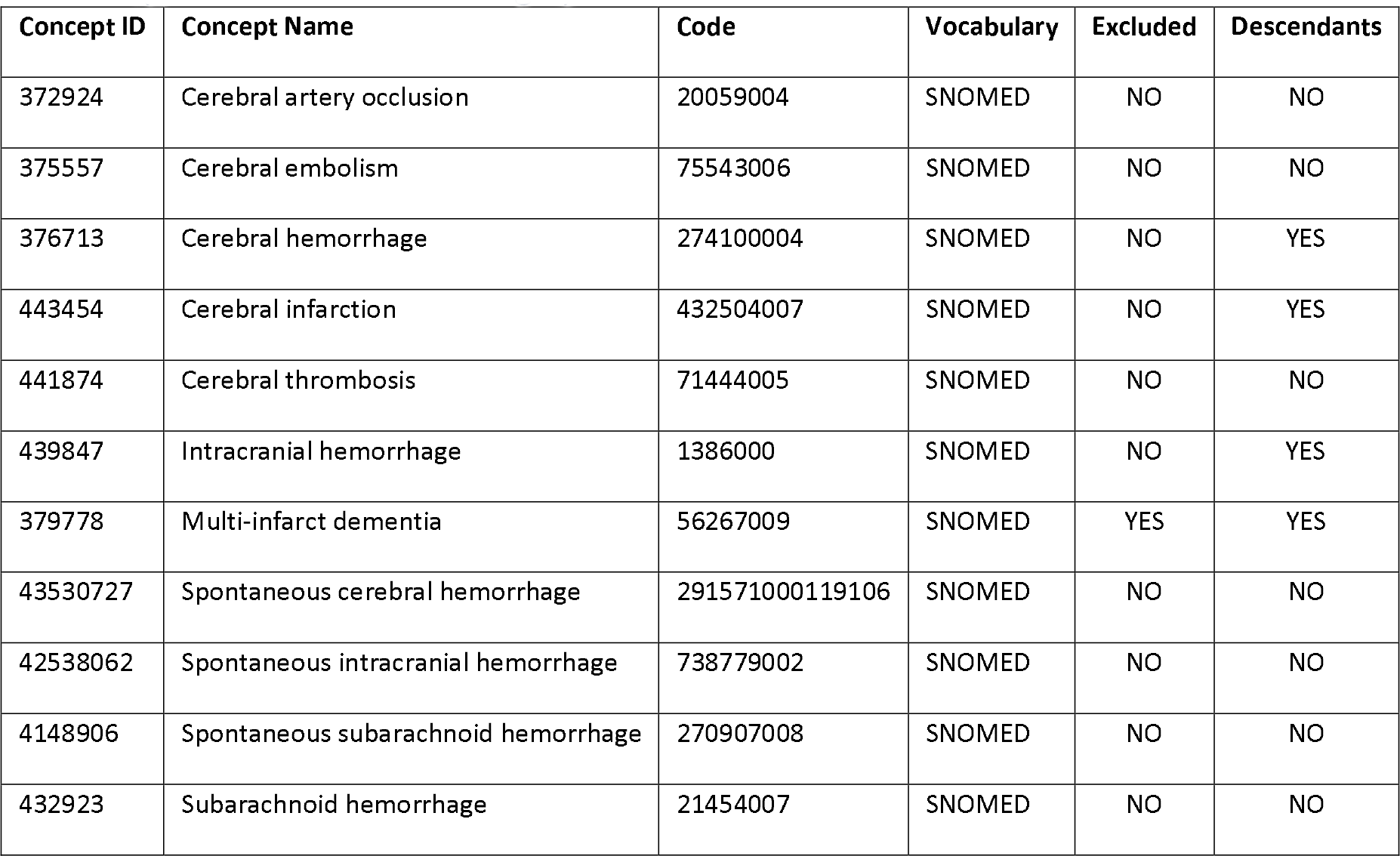

#### Inpatient or ER visit

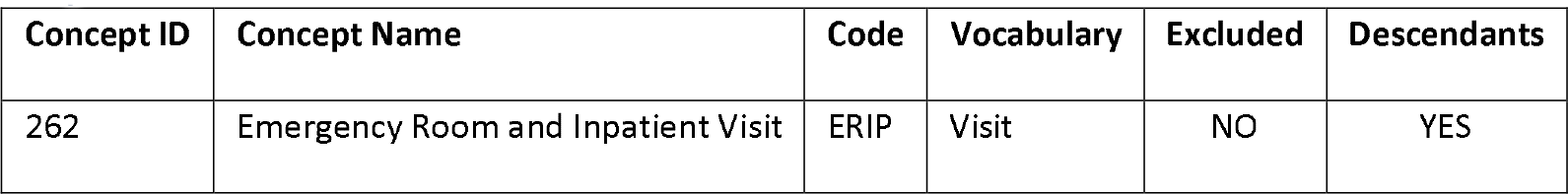
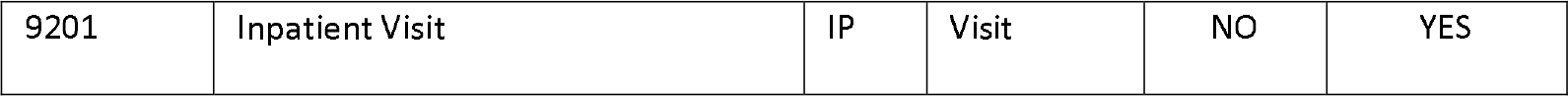

### Appendix 2

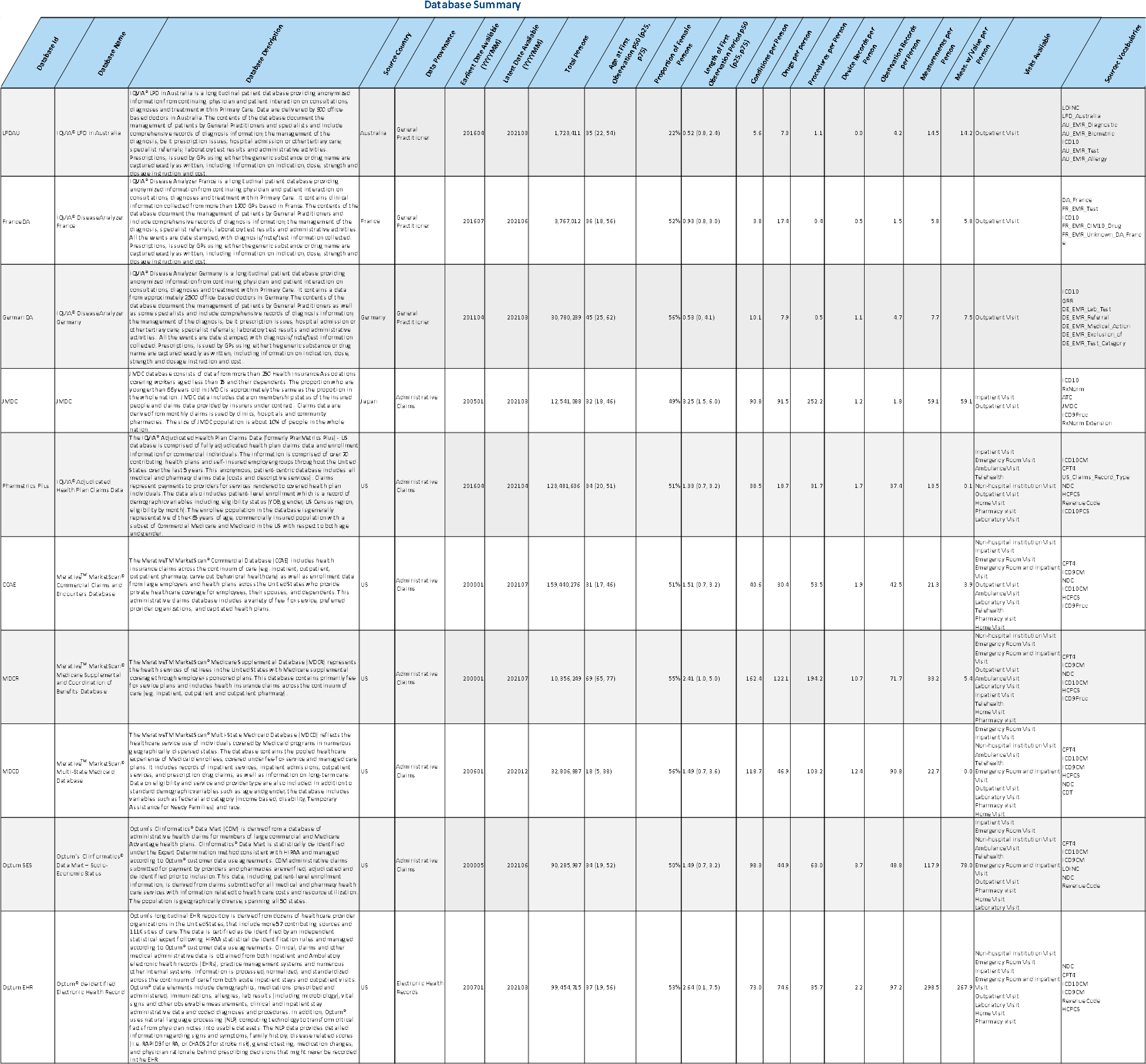

