## Appendix 1 for "CohortDiagnostics: phenotype evaluation across a network of observational data sources using population-level characterization"

**Appendix 2**

**Systemic lupus erythematosus indexed on signs, symptoms, treatment, or diagnosis (FP)**

**Human Readable Cohort Definition**

**Cohort Entry Events**

People enter the cohort when observing any of the following:

1. condition occurrences of ‘SLE or signs and symptoms suggestive of SLE’; having at least 1 condition occurrence of ‘Systemic lupus erythematosus (SLE)’ for the first time in the person’s history, starting between 0 days before and 90 days after ‘SLE or signs and symptoms suggestive of SLE’ start date.
2. drug exposures of ‘SLE treatments’; having at least 1 condition occurrence of ‘Systemic lupus erythematosus (SLE)’ for the first time in the person’s history, starting between 0 days before and 90 days after ‘SLE treatments’ start date.

Limit cohort entry events to the earliest event per person.

**Cohort Exit**

The person exits the cohort at the end of continuous observation.

**Cohort Eras**

Entry events will be combined into cohort eras if they are within 0 days of each other.

**Concept Sets:**

**Systemic lupus erythematosus (SLE)**

| **Concept ID** | **Concept Name** | **Code** | **Vocabulary** | **Excluded** | **Descendants** | **Mapped** |
| --- | --- | --- | --- | --- | --- | --- |
| 255891 | Lupus erythematosus | 200936003 | SNOMED | NO | YES | NO |
| 4300204 | Systemic lupus erythematosus-associated antiphospholipid syndrome | 402865003 | SNOMED | NO | YES | NO |
| 4319305 | Rash of systemic lupus erythematosus | 95332009 | SNOMED | NO | YES | NO |
| 4145240 | Renal tubulo-interstitial disorder in systemic lupus erythematosus | 307755009 | SNOMED | NO | YES | NO |
| 37016279 | Glomerular disease due to systemic lupus erythematosus | 308751000119106 | SNOMED | NO | YES | NO |
| 46273369 | Endocarditis due to systemic lupus erythematosus | 72181000119109 | SNOMED | NO | YES | NO |

**SLE treatments**

| **Concept ID** | **Concept Name** | **Code** | **Vocabulary** | **Excluded** | **Descendants** | **Mapped** |
| --- | --- | --- | --- | --- | --- | --- |
| 1777087 | hydroxychloroquine | 5521 | RxNorm | NO | YES | NO |
| 1551099 | prednisone | 8640 | RxNorm | NO | YES | NO |
| 1506270 | methylprednisolone | 6902 | RxNorm | NO | YES | NO |
| 1550557 | prednisolone | 8638 | RxNorm | NO | YES | NO |
| 1305058 | methotrexate | 6851 | RxNorm | NO | YES | NO |
| 19014878 | azathioprine | 1256 | RxNorm | NO | YES | NO |
| 40236987 | belimumab | 1092437 | RxNorm | NO | YES | NO |
| 1101898 | leflunomide | 27169 | RxNorm | NO | YES | NO |
| 19003999 | mycophenolate mofetil | 68149 | RxNorm | NO | YES | NO |

**SLE or signs and symptoms suggestive of SLE**

| **Concept ID** | **Concept Name** | **Code** | **Vocabulary** | **Excluded** | **Descendants** | **Mapped** |
| --- | --- | --- | --- | --- | --- | --- |
| 255891 | Lupus erythematosus | 200936003 | SNOMED | NO | YES | NO |
| 4300204 | Systemic lupus erythematosus-associated antiphospholipid syndrome | 402865003 | SNOMED | NO | YES | NO |
| 4319305 | Rash of systemic lupus erythematosus | 95332009 | SNOMED | NO | YES | NO |
| 4145240 | Renal tubulo-interstitial disorder in systemic lupus erythematosus | 307755009 | SNOMED | NO | YES | NO |
| 37016279 | Glomerular disease due to systemic lupus erythematosus | 308751000119106 | SNOMED | NO | YES | NO |
| 46273369 | Endocarditis due to systemic lupus erythematosus | 72181000119109 | SNOMED | NO | YES | NO |
| 439777 | Anemia | 271737000 | SNOMED | NO | NO | NO |
| 140214 | Eruption | 271807003 | SNOMED | NO | NO | NO |
| 45766714 | Inflammatory dermatosis | 703938007 | SNOMED | NO | NO | NO |
| 74125 | Inflammatory polyarthropathy | 417373000 | SNOMED | NO | NO | NO |
| 77074 | Joint pain | 57676002 | SNOMED | NO | NO | NO |
| 194133 | Low back pain | 279039007 | SNOMED | NO | NO | NO |
| 4272240 | Malaise | 367391008 | SNOMED | NO | NO | NO |
| 78517 | Multiple joint pain | 35678005 | SNOMED | NO | NO | NO |
| 138525 | Pain in limb | 90834002 | SNOMED | NO | NO | NO |
| 80809 | Rheumatoid arthritis | 69896004 | SNOMED | NO | NO | NO |

**Alzheimer’s disease (Simple)**

**Human Readable Cohort Definition**

**Cohort Entry Events**

People enter the cohort when observing any of the following:

1. condition occurrence of ‘Alzheimer’s disease’ for the first time in the person’s history.

Limit cohort entry events to the earliest event per person.

**Cohort Exit**

The person exits the cohort at the end of continuous observation.

**Cohort Eras**

Entry events will be combined into cohort eras if they are within 0 days of each other.

**Concept Sets:**

**Alzheimer’s disease**

| **Concept ID** | **Concept Name** | **Code** | **Vocabulary** | **Excluded** | **Descendants** | **Mapped** |
| --- | --- | --- | --- | --- | --- | --- |
| 378419 | Alzheimer’s disease | 26929004 | SNOMED | NO | YES | NO |

### Alzheimer's disease (based on Imfeld, 2013)

#### Human Readable Cohort Definition

| **Concept ID** | **Concept Name** | **Code** | **Vocabulary** | **Excluded** | **Descendants** |
| --- | --- | --- | --- | --- | --- |
| 378419 | Alzheimer's disease | 26929004 | SNOMED | NO | YES |

##### Dementia

| **Concept ID** | **Concept Name** | **Code** | **Vocabulary** | **Excluded** | **Descendants** |
| --- | --- | --- | --- | --- | --- |
| 37312036 | Aggression due to dementia | 788861009 | SNOMED | NO | YES |
| 37312035 | Agitation due to dementia | 788862002 | SNOMED | NO | YES |
| 4041685 | Amyotrophic lateral sclerosis with dementia | 230258005 | SNOMED | NO | YES |
| 37312031 | Anxiety due to dementia | 788866004 | SNOMED | NO | YES |
| 37312030 | Apathetic behavior due to dementia | 788867008 | SNOMED | NO | YES |
| 35608576 | Behavioral and psychological symptoms of dementia | 10171000132106 | SNOMED | NO | YES |
| 4092747 | Cerebral degeneration presenting primarily with dementia | 279982005 | SNOMED | NO | YES |
| 4182210 | Dementia | 52448006 | SNOMED | NO | YES |
| 37116464 | Dementia caused by heavy metal exposure | 733184002 | SNOMED | YES | NO |
| 37017549 | Dementia co-occurrent with human immunodeficiency virus infection | 713844000 | SNOMED | YES | NO |
| 4244346 | Dialysis dementia | 9345005 | SNOMED | YES | NO |
| 37311665 | Disinhibited behavior due to dementia | 789170003 | SNOMED | NO | YES |
| 4043378 | Frontotemporal dementia | 230270009 | SNOMED | NO | YES |
| 45765480 | Frontotemporal dementia with parkinsonism-17 | 702429008 | SNOMED | NO | YES |
| 377788 | General paresis - neurosyphilis | 51928006 | SNOMED | YES | NO |
| 45765477 | GRN-related frontotemporal dementia | 702426001 | SNOMED | NO | YES |
| 4059191 | H/O: dementia | 161465002 | SNOMED | NO | YES |
| 372610 | Postconcussion syndrome | 40425004 | SNOMED | YES | NO |
| 37017247 | Presenile dementia co-occurrent with human immunodeficiency virus infection | 713488003 | SNOMED | YES | NO |
| 37311890 | Psychological symptom due to dementia | 789011007 | SNOMED | NO | YES |
| 37312577 | Wandering due to dementia | 789062005 | SNOMED | NO | YES |

##### Prescription for an Alzheimers disease drug

| **Concept ID** | **Concept Name** | **Code** | **Vocabulary** | **Excluded** | **Descendants** |
| --- | --- | --- | --- | --- | --- |
| 715997 | donepezil | 135447 | RxNorm | NO | YES |
| 757627 | galantamine | 4637 | RxNorm | NO | YES |
| 701322 | memantine | 6719 | RxNorm | NO | YES |
| 733523 | rivastigmine | 183379 | RxNorm | NO | YES |
| 836654 | tacrine | 10318 | RxNorm | NO | YES |

##### Specific dementia test

| **Concept ID** | **Concept Name** | **Code** | **Vocabulary** | **Excluded** | **Descendants** |
| --- | --- | --- | --- | --- | --- |
| 4169175 | Mini-mental state examination | 273617000 | SNOMED | NO | YES |
| 40491929 | Mini-mental state examination score | 447316007 | SNOMED | NO | YES |
| 40490379 | Assessment using mini-mental state examination | 446971008 | SNOMED | NO | YES |
| 4167593 | Abbreviated Mental Test | 273255001 | SNOMED | NO | YES |
| 4013636 | Magnetic resonance imaging | 113091000 | SNOMED | NO | YES |
| 4125350 | CT of head | 303653007 | SNOMED | NO | YES |
| 4019823 | Single photon emission computerized tomography | 105371005 | SNOMED | NO | YES |

##### Dementia symptoms

| **Concept ID** | **Concept Name** | **Code** | **Vocabulary** | **Excluded** | **Descendants** |
| --- | --- | --- | --- | --- | --- |
| 4304008 | Memory impairment | 386807006 | SNOMED | NO | YES |
| 440424 | Aphasia | 87486003 | SNOMED | NO | YES |
| 132342 | Apraxia | 68345001 | SNOMED | NO | YES |
| 4173136 | Agnosia | 42341009 | SNOMED | NO | YES |
| 4024716 | Aphasia, agnosia, dyslexia AND/OR apraxia | 106169008 | SNOMED | NO | YES |

##### Other specific dementia diagnosis (e.g., VD, Pick’s disease, or Lewy body dementia [LBD])

| **Concept ID** | **Concept Name** | **Code** | **Vocabulary** | **Excluded** | **Descendants** |
| --- | --- | --- | --- | --- | --- |
| 443605 | Vascular dementia | 429998004 | SNOMED | NO | YES |
| 44782710 | Dementia due to Pick's disease | 21921000119103 | SNOMED | NO | YES |
| 380701 | Diffuse Lewy body disease | 80098002 | SNOMED | NO | YES |

##### Stroke (ischemic or hemorrhagic)

| **Concept ID** | **Concept Name** | **Code** | **Vocabulary** | **Excluded** | **Descendants** |
| --- | --- | --- | --- | --- | --- |
| 372924 | Cerebral artery occlusion | 20059004 | SNOMED | NO | NO |
| 375557 | Cerebral embolism | 75543006 | SNOMED | NO | NO |
| 376713 | Cerebral hemorrhage | 274100004 | SNOMED | NO | YES |
| 443454 | Cerebral infarction | 432504007 | SNOMED | NO | YES |
| 441874 | Cerebral thrombosis | 71444005 | SNOMED | NO | NO |
| 439847 | Intracranial hemorrhage | 1386000 | SNOMED | NO | YES |
| 379778 | Multi-infarct dementia | 56267009 | SNOMED | YES | YES |
| 43530727 | Spontaneous cerebral hemorrhage | 291571000119106 | SNOMED | NO | NO |
| 42538062 | Spontaneous intracranial hemorrhage | 738779002 | SNOMED | NO | NO |
| 4148906 | Spontaneous subarachnoid hemorrhage | 270907008 | SNOMED | NO | NO |
| 432923 | Subarachnoid hemorrhage | 21454007 | SNOMED | NO | NO |

##### Inpatient or ER visit

| **Concept ID** | **Concept Name** | **Code** | **Vocabulary** | **Excluded** | **Descendants** |
| --- | --- | --- | --- | --- | --- |
| 262 | Emergency Room and Inpatient Visit | ERIP | Visit | NO | YES |
| 9201 | Inpatient Visit | IP | Visit | NO | YES |
